# Within-host virus evolution during the extended treatment of RSV infection with mutagenic drugs

**DOI:** 10.1101/2022.09.02.22279474

**Authors:** Christopher J. R. Illingworth, Alexandra Y. Kreins, Adriana Margarit-Soler, Tim Best, Patricia Dyal, Giovanna Lucchini, Kanchan Rao, Rachel Williams, Austen Worth, Judith Breuer

**Author notes:** Authors contributed equally (Christopher J. R. Illingworth, Alexandra Y. Kreins).

## Abstract

Antiviral drugs causing viral mutagenesis have shown value against a broad range of RNA viruses causing respiratory illnesses. While drug-induced accumulation of mutations generally decreases viral fitness, the potential for mutagenesis to generate escape variants is unknown and concerns have been raised about adaptive evolution promoting drug-resistance. We report prolonged treatment of a life-threatening RSV infection with a combination of two viral RNA-dependent RNA polymerase (RdRp) inhibitors, ribavirin and favipiravir, in a child with severe combined immunodeficiency undergoing haematopoietic stem cell transplantation. Viral deep sequencing of longitudinally collected RSV samples determined that ribavirin caused a 3-fold increase in the viral mutation rate. There was no synergistic effect upon addition of favipiravir. Viral load remained unchanged throughout antiviral treatment, but genomic modelling predicted loss of viral fitness secondary to drug-induced mutagenesis. The viral changes coincided with stabilisation of the patient’s clinical condition. In the absence of viral clearance, adaptive evolution occurred on a complex fitness landscape, leading to increased population diversity at the haplotype level. The evolutionary consequences of using mutagenic antiviral drugs are likely to be hard to predict, but in this example within-host virus evolution under extended treatment with mutagenic drugs resulted in an overall loss of viral fitness due to deleterious mutations accumulating faster than could be outweighed by positive selection. These genomic findings occurred in tandem with evidence of clinical improvement and are potentially associated.

## Introduction

Respiratory syncytial virus (RSV) is the primary cause of hospitalization with acute lower respiratory infection (LRTI) among children, with approximately 3.2 million hospitalisations worldwide in 2015 (Shi et al., 2017). Public health measures taken during the COVID-19 pandemic have reduced the incidence of RSV infection (Britton et al., 2020; van Summeren et al., 2021), but concerns remain that this reduction of cases may have led to the accumulation of an ‘immunity debt’ (Hatter et al., 2021), generating the potential for a future RSV epidemic of increased severity. There currently is no licensed vaccine against RSV, despite candidates making it to Phase 3 clinical trials (Ginsburg and Srikantiah, 2021; Madhi et al., 2020). Monoclonal antibodies have been used for so-called passive immunisation and have shown some potential to reduce hospitalisation of premature infants predisposed to cardio-respiratory conditions (Morris et al., 2009; O’Brien et al., 2015), with palivizumab being licensed for use (Mejias and Ramilo, 2008; Wegzyn et al., 2014). Although no randomised trials have been conducted, palivizumab prophylaxis has been recommended in immunosuppressed patients below the age of two (Hirsch et al., 2013). It is recognised that vulnerable patients who are at risk of life-threatening respiratory RSV infections also urgently require access to effective antiviral treatments. For example, RSV+ LRTIs are associated with 30% excess mortality in paediatric recipients of haematopoietic stem cell transplantation (HSCT) (Adams et al., 1999). For this reason it has been recommended to delay HSCT to allow for LTRI treatment, most commonly with one antiviral, ribavirin, in order to reduce HSCT-related mortality (Ottaviano et al., 2020).

Ribavirin is licensed for treatment of RSV infection in children with severe bronchiolitis especially when they have other underlying illness (Hall et al., 1983; Turner et al., 2014), and has successfully been used for pre-emptive treatment of RSV infections in these patients (Adams et al., 1999; Sparrelid et al., 1997). However, its use has been limited by its cost and by concerns around drug toxicity (Domachowske et al., 2021), and its efficacy as a treatment for established RSV infection, especially in immunocompromised patients, has not been established. The drug is an inhibitor of the RNA dependent RNA polymerase (RdRp) of RNA viruses, such as RSV, inducing C to U and G to A mutations in the viral genome during replication (Vo et al., 2003). RdRp inhibitors have the potential for broad-spectrum antiviral activity and have been repurposed for use against a growing number of RNA viruses. Favipiravir, another RdRp inhibitor, has been licensed in Japan for the treatment of influenza and its mutagenic effect has been demonstrated with accumulation of signature C to U and G to A mutations (Goldhill et al., 2019). Favipiravir treatment efficacy against influenza has not convincingly been demonstrated in trials (Furuta et al., 2017; Hayden et al., 2022; Wang et al., 2020a), but good outcomes have been reported for combination therapy (Wang et al., 2020b). For example, we previously reported the successful use of favipiravir in combination with zanamivir, a nucleoside inhibitor, against chronic influenza B infection in an immunocompromised child (Lumby et al., 2020b). This illustrates that RdRp inhibitors can be repurposed on a compassionate basis as a bridging therapy to corrective cellular therapy in immunodeficient patients with life-threatening RNA virus infections.

Further interest in studying mutagenic drugs has arisen during the SARS-CoV-2 pandemic (Brown et al., 2021). Animal studies showed that favipiravir-induced mutagenesis resulted in loss of SARS-CoV-2 infectivity (Kaptein et al., 2020). Combination treatment with molnupiravir, another RdRp inhibitor which induces viral mutagenesis, further amplified this (Abdelnabi et al., 2021). Molnupiravir has been shown to reduce the risk of hospitalisation or death if given early in SARS-CoV-2 infection (Gordon et al., 2021; Jayk Bernal et al., 2022; Kabinger et al., 2021), but concerns have been raised about the potential for mutagenesis to have harmful side-effects. Negative outcomes could include the induction of mutations in the cells of the person treated (Swanstrom and Schinazi, 2022), or the promotion of antiviral resistance, such as that observed to favipiravir in influenza (Goldhill et al., 2018). The evolution of SARS-CoV-2 in immunocompromised hosts in some ways reflects patterns seen in the emergence of novel viral variants (Harari et al., 2022); if antivirals promoted harmful patterns of evolution and were disproportionately used among this cohort that might have global consequences. Before widespread repurposing of RdRp inhibitors, these concerns need to be addressed.

We here report treatment outcomes for one paediatric patient with severe combined immunodeficiency (SCID) and RSV pneumonitis. Before proceeding with HSCT, the patient was treated with ribavirin, initially in combination with nitazoxanide, which has broad spectrum antiviral effects against RNA viruses (Blum et al., 2021; Jasenosky et al., 2019), and subsequently in combination with favipiravir, for which *in vitro* activity specifically against RSV has been reported (Furuta et al., 2002). The combination of ribavirin and favipiravir has been shown to be synergistic against influenza *in vitro* through distinct modes of action on its polymerase (Vanderlinden et al., 2016). In contrast to short trial protocols for favipiravir and/or molnupiravir in patients with RNA virus infections, including influenza, ebola and SARS-CoV-2 (Brown et al., 2021; Guedj et al., 2018; Hayden et al., 2022; Jayk Bernal et al., 2022), antiviral treatment in this SCID patient was administered over an extended period of 2 months during which time the patient received a cord blood HSCT from a matched unrelated donor. While the patient’s respiratory function improved during the antiviral treatment, she did not clear the infection. Taking advantage of the persistent viral load (VL) during prolonged antiviral treatment in this patient, we used viral genome sequencing data alongside population genetic modelling to investigate the impact of drug-induced mutagenesis on the RSV population. We investigated how mutagenic drugs shaped the viral population, taking a genomic perspective on the question of whether drug treatment contributed to clinical improvement.

## Results

### Treatment of a RSV+ LRTI with RdRp inhibitors in a SCID patient awaiting HSCT

The patient under investigation was diagnosed during infancy with T-B+NK+SCID due to IL-7Ra deficiency following recurrent upper respiratory tract infections. Two months following diagnosis she underwent a first HSCT procedure. She displayed early donor engraftment, but this was not sustained over time with a progressive decrease in donor chimerism. RSV was first detected in the nasopharyngeal aspirate (NPA) 6 months after transplantation with a Ct of 22 and continued to be persistently detected in repeat NPAs. The patient also developed adenoviraemia and was referred for a second HSCT procedure. In order to reduce the risk of post-HSCT mortality in the context of an RSV+ LRTI, treatment was initiated with nebulised ribavirin (6g daily) in combination with nitazoxanide (200mg 12-hourly), and weekly intravenous immunoglobulin therapy. After 4 weeks of this treatment regimen, there was no VL reduction (Figure 1). Nitazoxanide was discontinued and favipiravir (200mg 8-hourly) was started in addition to ribavirin. VL remained unchanged (Figure 1). 12 days later she underwent a second HSCT procedure with cord blood stem cells from a matched unrelated donor; for the purposes of our study we denote the date of this transplant as day 0. From a respiratory point of view, she remained asymptomatic despite receiving reduced intensity conditioning with targeted busulfan and serotherapy with ATG pre-HSCT, as well as immunosuppression with ciclosporin and mycophenolate mofetil for graft-versus-host-disease (GVHD) prophylaxis post-HSCT. Favipiravir was discontinued 18 days after HSCT. Ribavirin, which after HSCT was changed from inhalation to oral administration (50mg 8-hourly), was stopped 11 days later. One month after this second HSCT, mixed donor chimerism was first detected in whole blood and granulocytes, in absence of lymphoid engraftment. Donor chimerism in the lymphoid lineages increased with progressive reduction and subsequent cessation of the immunosuppressive GVHD prophylaxis. As of 6 months post-HSCT the patient displayed improving immune reconstitution with full donor engraftment and RSV was no longer detected in her NPA (Figure 1).

**Figure 1:**
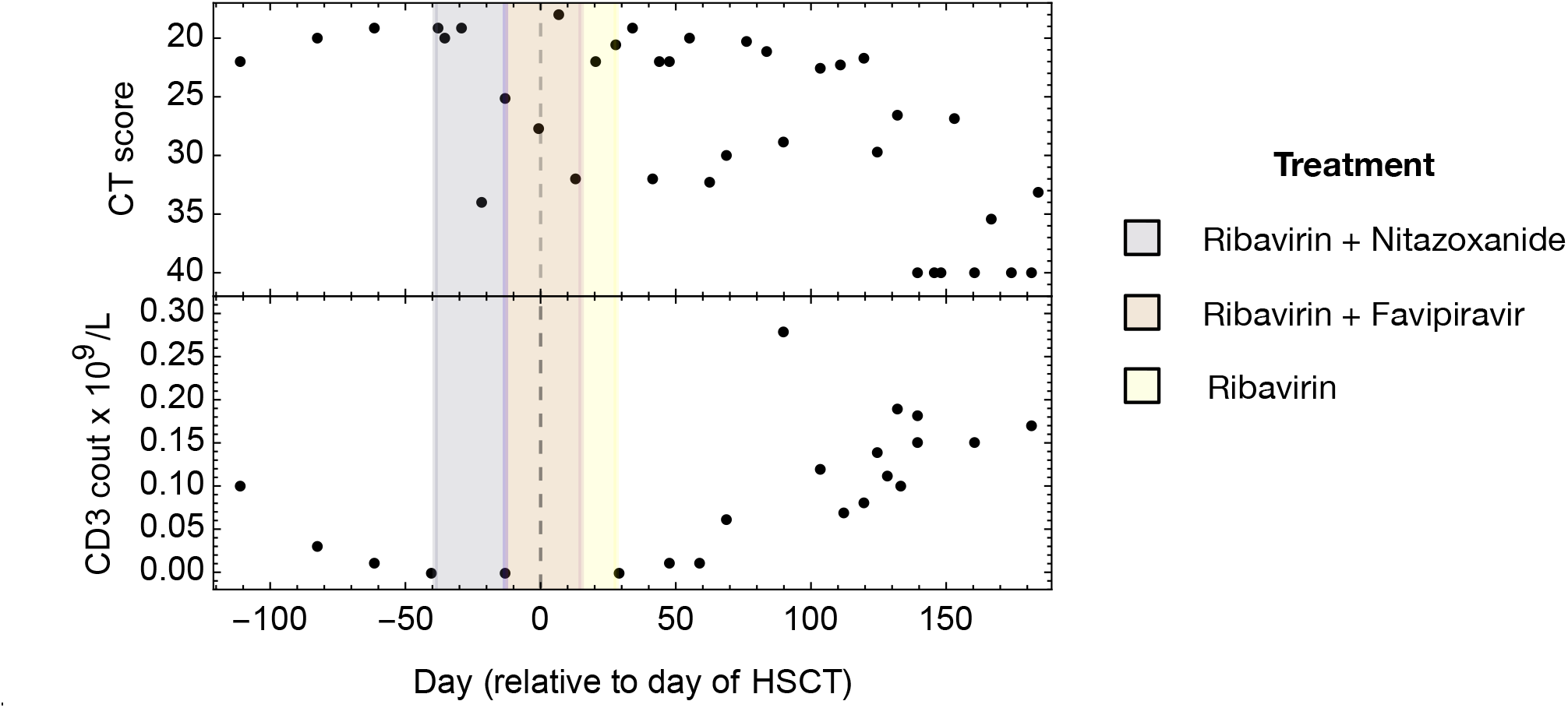
Changes in viral load and CD3 cell count during RSV infection within an immunocompromised child. Recovery from infection corresponded to a recovery in T-cell immunity. Black dots show CT scores and cell count data. A vertical black dashed line shows the time at which the child received an HSCT. Shading shows times of treatment with ribavirin, nitazoxanide, and favipiravir.

In summary, we report clinical stabilisation in an immunodeficient patient with RSV+ LRTI undergoing HSCT during antiviral treatment with ribavirin in combination with nitazoxanide or favipiravir, without viral clearance. Notwithstanding the day-to-day variability of Ct values in NPA samples, VL overall remained unchanged throughout the period of treatment, with no apparent impact by any of the three drugs used.

### Genomic consequences of treatment with RdRp inhibitors

Next-generation sequencing was performed for RSV isolated from 10 NPA samples, including 3 samples collected prior to treatment initiation, 2 collected during the initial treatment regimen with ribavirin and nitazoxanide, 4 during treatment with ribavirin and favipiravir, and 1 during subsequent treatment with ribavirin alone (Figure 2). The results showed substantially increased mutational load in the viral population during treatment despite no fall in VL. Mutational load, equal to the sum of minor allele frequencies across the genome (Lumby et al., 2020a), describes the size of the burden carried by viruses resulting from deleterious mutations in the genome, and was calculated for each sample. Fitting a model to these data suggested that treatment with ribavirin led to a roughly three-fold increase in the viral mutation rate (Figure 2A); a more complex model which allowed for a distinct mutation rate upon the addition of favipiravir did not give a significantly better fit to the data (Figure 2B). Our model of mutational load makes the assumption of a large effective population size for the virus (Lumby et al., 2020a); an analysis of changes in the viral population over time found negligible signal of directed evolutionary change in the viral population, compatible with a slow rate of genetic drift and therefore with this assumption (Figure 2S1).

**Figure 2:**
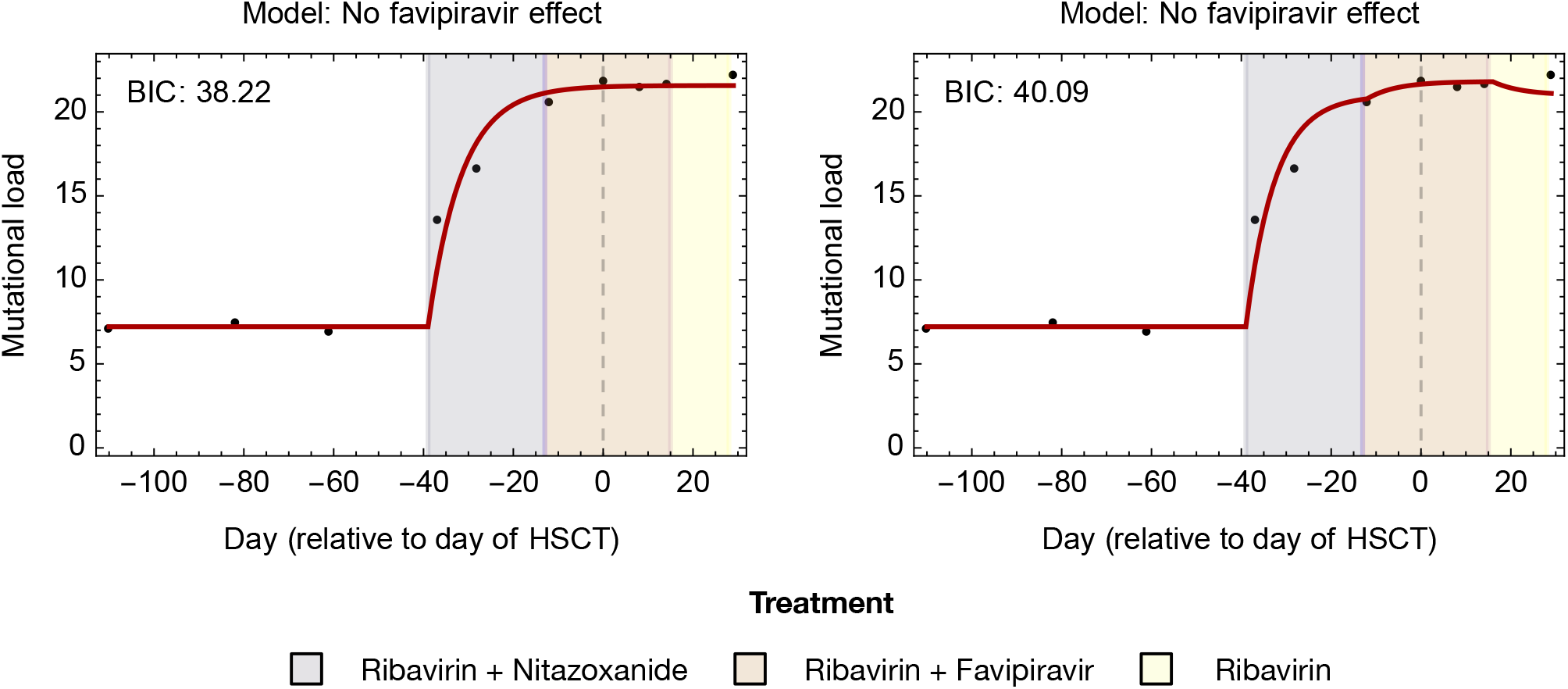
Models describing changes in mutational load over time following treatment. Black dots show the measured mutational load in the viral population in samples collected over time. Red lines show the maximum likelihood fit of each model to the data. Models fitted to the data either allow or disallow favipiravir to have an independent effect on the viral mutation rate. A lower BIC indicates a better fit between the model and the data, accounting for the complexity of the model.

Changes in the mutational spectra of the viral population further confirmed the association of ribavirin with increased mutagenesis. C to U mutations increased during treatment, comprising a mean of 10.2% of mutational load in the first three samples, but a mean of 20.8% of load in the last three samples collected. Similarly, G to A mutations increased from a mean of 6.9% of mutational load in the first three samples collected to a mean of 11.4% across the last three samples (Figure 3).

**Figure 3:**
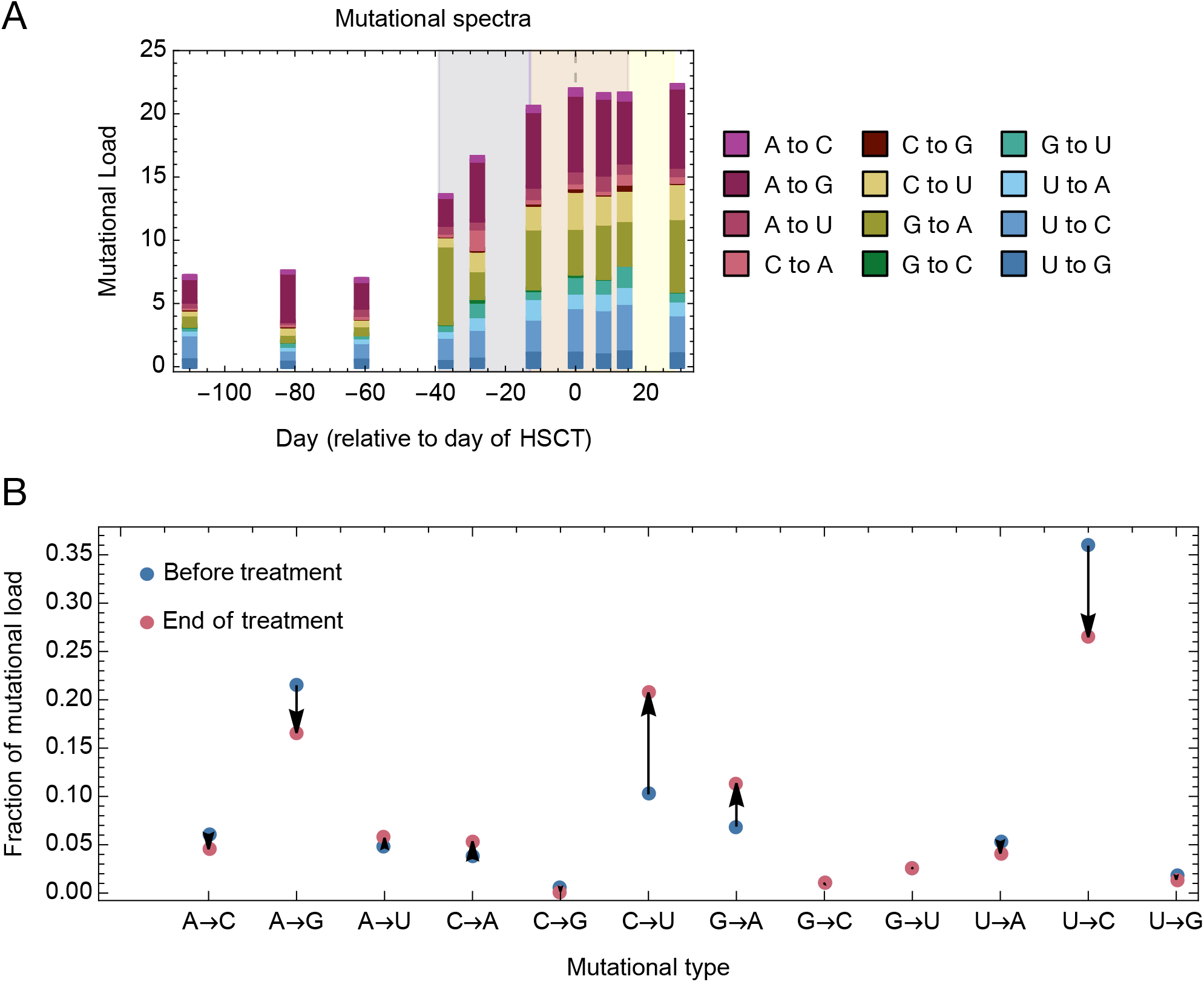
Mutational spectra as a function of mutational class. **A**. Breakdown of mutational load by mutational class. Background shading shows times of treatment with ribavirin, nitazoxanide, and favipiravir. **B**. Proportion of mutational load comprised of each mutational class in the first three (Before treatment) and last three (End of treatment) samples. Treatment increased the proportion of C to T and G to A mutations.

### Fitness costs of viral mutagenesis

While treatment did not clear viral infection, the inference of a change in the viral mutation rate suggests that ribavirin may have nonetheless reduced viral fitness, potentially contributing to the relatively benign clinical course of RSV infection in this patient undergoing HSCT. An increased mutation rate leads to each virus carrying an increased number of deleterious mutations, which inevitably decreases the fitness of the viral population. At the same time, a reduction in viral fitness may not be accompanied by a large decrease in VL, dependent upon the initial fecundity of the virus. Viral fecundity describes the mean number of cells that will be infected by the viruses produced by a single infected cell: If a reduction in fitness does not cause this statistic to fall below one, the viral population will not die out (Bull et al., 2007).

The precise decline in fitness in the RSV population was difficult to quantify, with our model containing some redundancy dependent upon the generation time of the viral population (Figure 4A). Assuming a generation time of between 6 and 48 hours (Baccam et al., 2006; Perelson et al., 1996) we calculated that ribavirin caused between a 38% and a 98% decrease in viral fitness, a longer generation time leading to a greater loss of fitness (Figure 4B, Figure 4S1). Across realistic potential values for viral fecundity prior to treatment (Baccam et al., 2006), and under a simple birth-death model of replication, such reductions in fitness are compatible with a failure to clear viral infection (Figure 4C,D). Given a long generation time, even a high pre-treatment fecundity would have been overcome by the resultant loss of fitness. However, at shorter generation times fitness may be lost without causing an observable signal in the viral load. The observed failure to clear viral infection suggests a generation time for RSV of less than 40 hours, but without further knowledge of the virus we are unable to conclude more than this.

**Figure 4:**
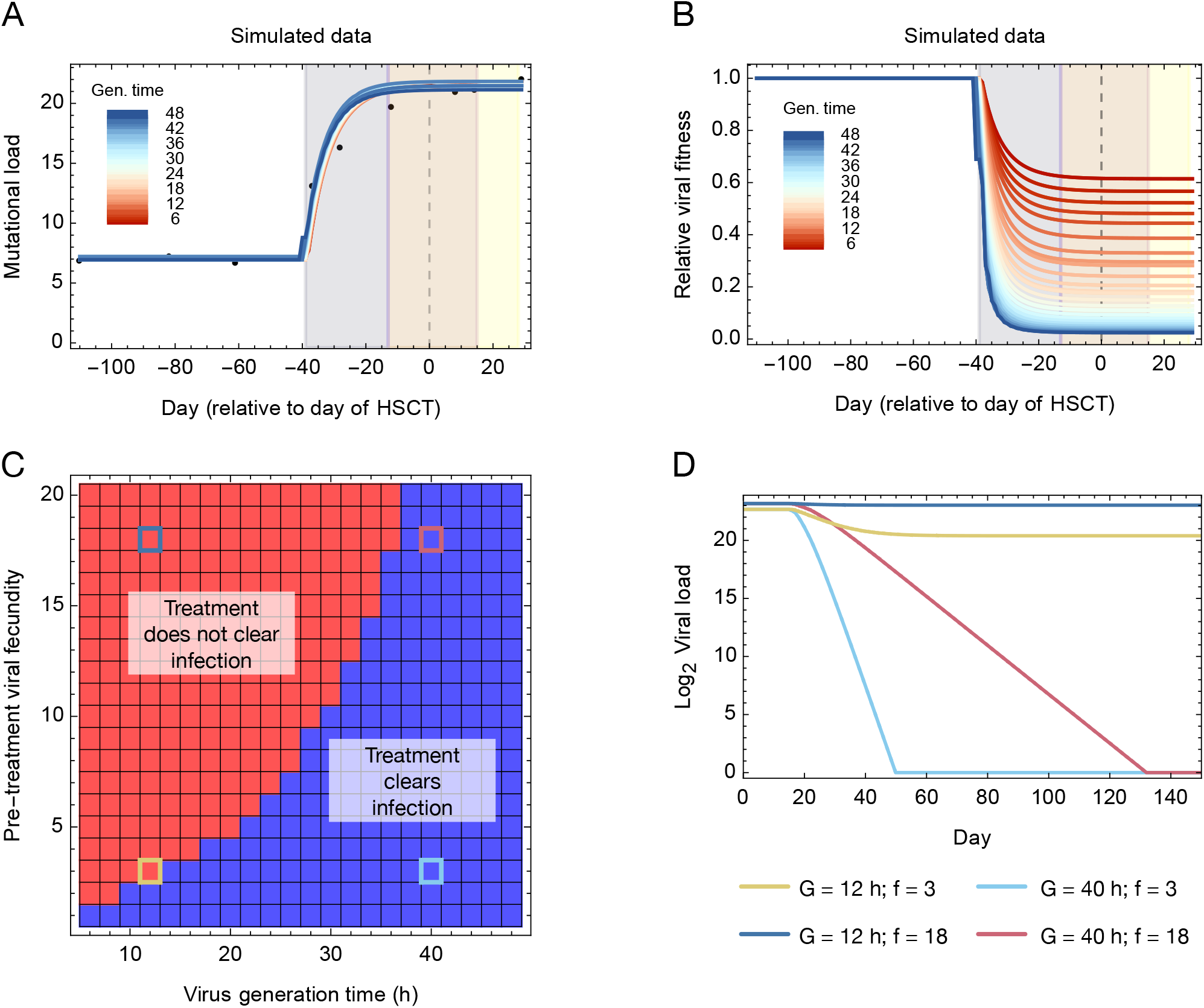
Effect of mutational load upon viral fitness. **A**. Near-identical fits to the mutational load data can be generated at different assumed viral generation times, with differences in the visualised curves arising only from interpolation effects. Background shading shows times of treatment with ribavirin, nitazoxanide, and favipiravir. **B**. Changes in the relative fitness of the virus are strongly dependent upon the assumed generation time. **C**. The fate of the viral population given treatment depends upon the assumed generation time and the fecundity of the viral population prior to treatment. Shading shows parameters which lead to the clearance of infection (blue) or the failure of treatment to clear infection (red), the latter corresponding to the clinical observations. Squares with highlighted borders correspond to specific sets of parameters chosen for further analysis. **D**. Expected changes in viral load given these specific parameters, generated by a simple birth-death model with a carrying capacity. Within the model, treatment begins on day 15.

### Limited adaptive evolution despite mutagenesis

We considered adaptive evolution within the viral population by studying variants observed at higher frequencies. Among 79 genetic variants observed at frequencies in excess of 5% in at least two samples (Supplementary Table 1), seven were identified as being potentially influenced by selection (Figure 5A). Of these, five were non-synonymous and two synonymous. Correlations between the frequencies of some of these variants were consistent with genetic hitchhiking (Figure 5S1). None of the potentially selected variants were associated with transcription, suggesting that they are unlikely to have influenced the interaction between RSV and either ribavirin or favipiravir.

**Figure 5:**
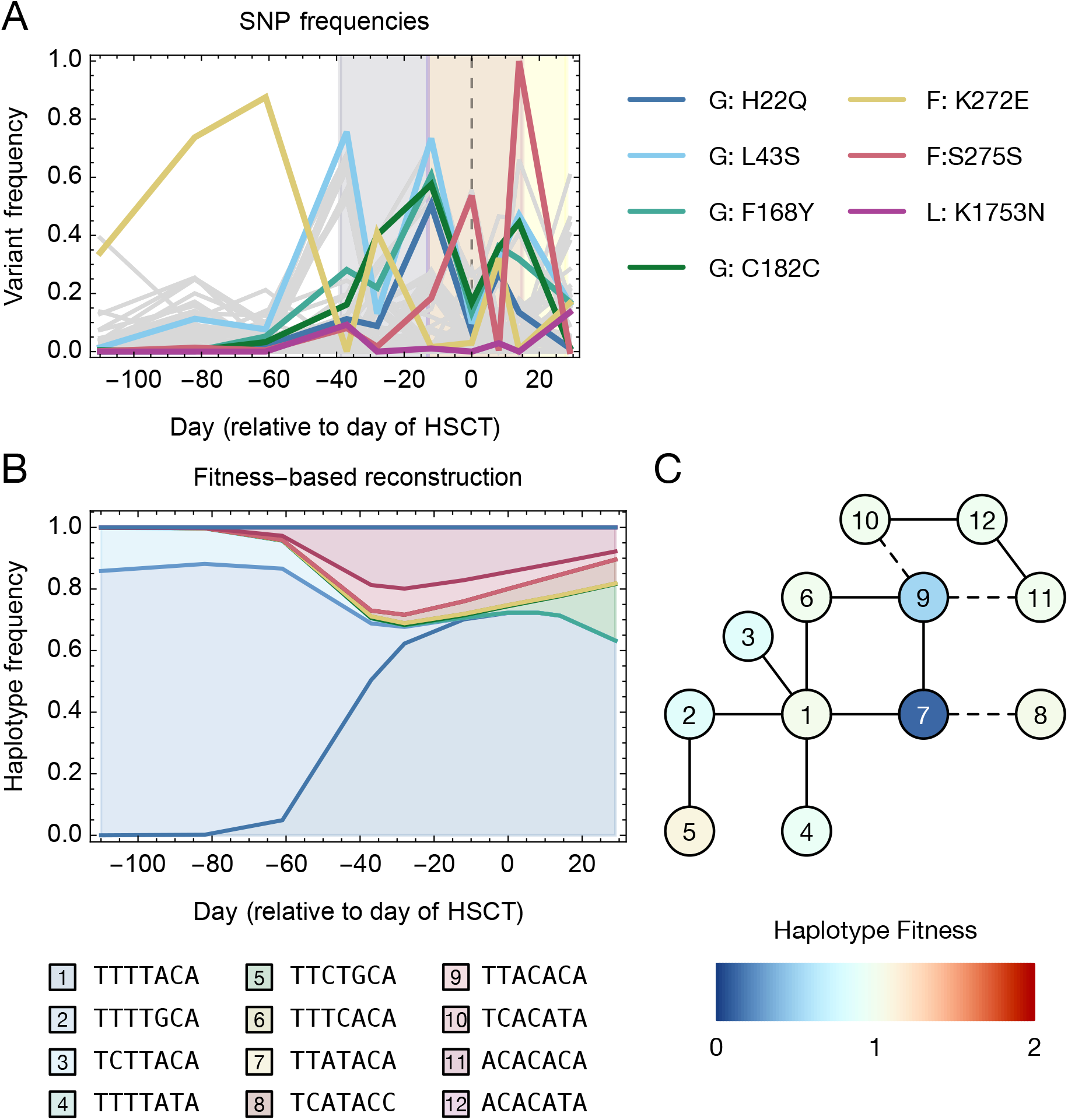
Within-host variation. **A**. Allele frequency trajectories of all variants that were observed at a frequency of at least 5% in at least two samples. Trajectories are shown in gray with potentially selected variants in colour. Background shading shows times of treatment with ribavirin, nitazoxanide, and favipiravir. **B**. Evolutionarily constrained haplotype reconstruction, in which each haplotype maintains a constant fitness over time. Not all of the haplotypes reach frequencies sufficient to be observed in the plot. **C**. Genetic relationships between haplotypes and inferred fitnesses. A solid line indicates haplotypes that are separated by a single mutation while a dashed line indicates haplotypes that are separated by two mutations.

Of the seven variants, the substitution F168Y in the attachment glycoprotein G is of particular interest. This variant arises in the central conserved region of G (McLellan et al., 2013); structural evidence showing the binding of this region by a neutralising antibody (Jones et al., 2018) highlights that the F168Y substitution would introduce a hydroxyl group into the binding interface (Figure 5S2), potentially leading to host immune escape. This could constitute a functional advantage for RSV. Of the other variants, the K272E substitution in the fusion glycoprotein has been identified as conferring resistance to the monoclonal antibody palivizumab in an animal system, the substitution being sited at the binding interface with this antibody (Swanson et al., 2011; Zhao et al., 2004). At the time of infection, the patient was receiving monthly palivizumab prophylaxis, which had been approved until the end of the RSV season. Upon cessation of palivizumab, the frequency of this variant decreased over time within the population, suggesting reversion to the population-wide consensus.

The escape of viruses from the deleterious effects of mutagenesis could occur either through the generation of strongly beneficial single mutations, or through the generation of high fitness combinations of mutations. An assessment of fitness at the level of viral haplotypes showed a slow process of within-host evolution, with minor gains in viral fitness. A process of haplotype reconstruction conducted at the seven potentially selected sites suggested that viral diversity at these sites could be explained in terms of twelve underlying viral haplotypes (Figure 5S3). Of note, haplotypes containing the F168Y substitution (having an A at the third nucleotide position) appear at higher frequencies following the use of ribavirin. However, these haplotypes were not inferred to have substantially higher fitness than other viral sequences. A second reconstruction, in which haplotype frequencies were constrained to follow an underlying evolutionary model, suggested that the fittest observed haplotype, which contained the L43S and F168Y substitutions in G, and the K1753N substitution in L, had a fitness just 26% greater than the mean fitness of the initial population, and just 5% greater than the mean fitness of the population at the time of the last sample (Figure 5B,C). Rather than a simple pattern of the virus gaining successive beneficial mutations, adaptation was multi-directional, with the haplotypes that comprised the final population (5, 8, 10, and 11) occurring in distinct parts of genotype space. In summary, despite prolonged treatment with effective mutagenic drugs, a raised mutation rate did not lead to the emergence of highly beneficial escape mutations.

### Adaptive evolution did not overcome the loss of fitness caused by mutagenesis

Adaptive evolution and the accumulation of mutational load occurred in parallel, with competition between haplotypes occurring while each haplotype gained deleterious mutations through an increased rate of viral mutation. Combining models showed that even under conservative assumptions the gain in viral fitness through adaptive evolution fell short of the corresponding loss of fitness acquired through mutational load (Figure 6). Within our reconstruction the different components of our model have differing complexity. Once the mutation rate is altered, under our model the loss of fitness due to viral mutagenesis is fixed. By contrast, the complexity of the landscape of viral adaptation leads to less uniform gains via adaptive evolution over time. Nevertheless, in this case of RSV infection, the costs of mutational load outweighed the benefits of adaptive evolution. Extended treatment with an effective mutagenic drug did not lead to the generation of beneficial variants sufficient to escape the effects of mutational load.

**Figure 6:**
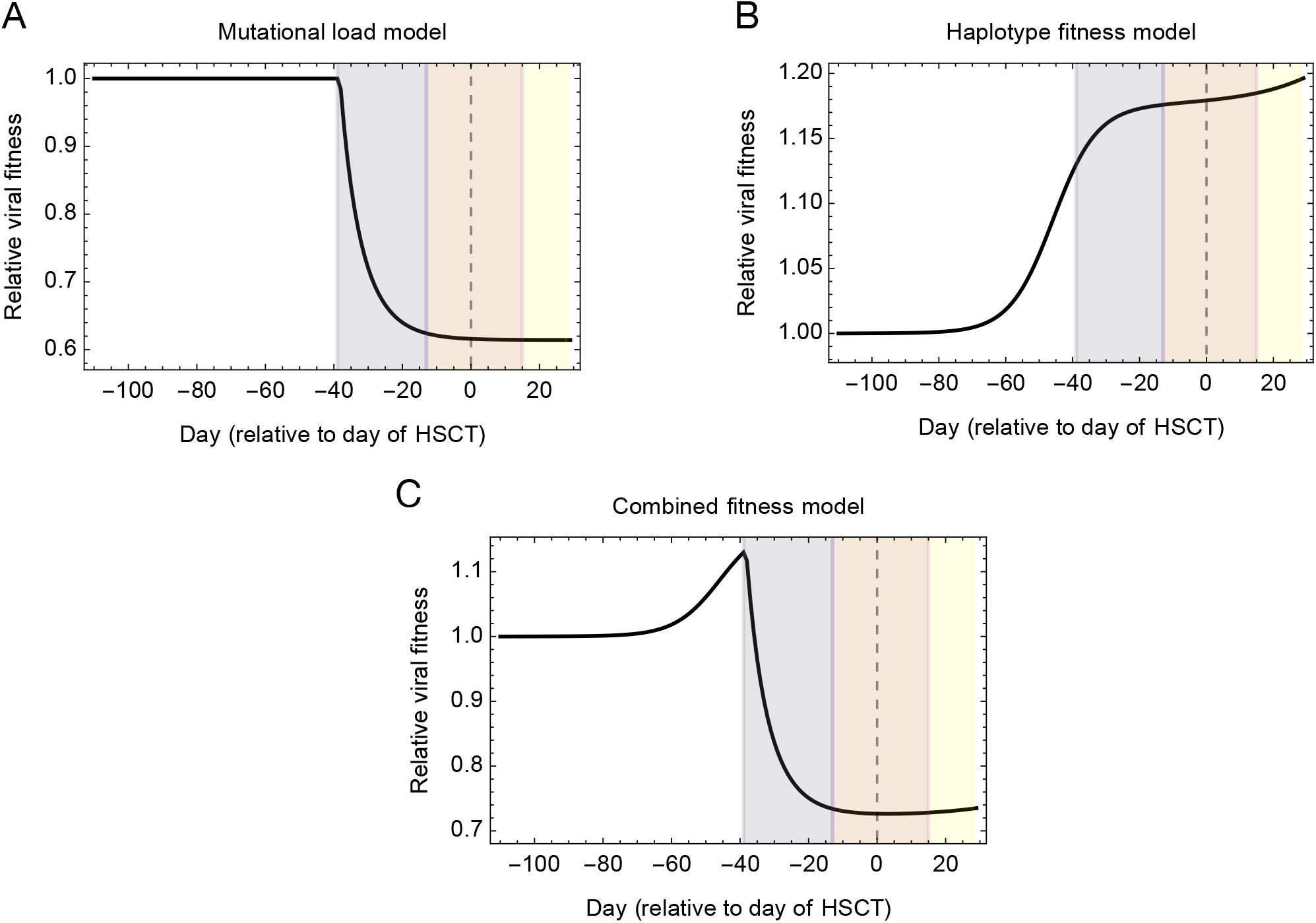
Reconstruction of viral fitness over time. **A**. Changes in the relative fitness of the viral population inferred according to our model of mutational load, under the conservative assumption of a viral generation time of 6 hours. Background shading shows times of treatment with ribavirin, nitazoxanide, and favipiravir. **B**. Changes in the relative fitness of the viral population inferred under our model of haplotype-based evolution. **C**. A combined model of fitness combines fitness components from both models in a multiplicative manner, showing the gain of fitness due to adaptive evolution in combination with the inferred loss of fitness through the gain of deleterious mutations arising through viral mutagenesis. We note that, even under a conservative model of the effects of viral load, adaptive evolution does not compensate for the increase in mutational load.

## Discussion

We have here described a case of long-term RSV infection in a severely immunocompromised patient, in whom a benign clinical course followed HSCT until eventual clearance of virus occurred with donor T-cell engraftment and reconstitution of T-cell immunity. During the course of infection the patient received extended treatment with the mutagenic antiviral drugs ribavirin and favipiravir. Analysis of changes in viral genome sequence data over time provided insights into the evolutionary response of the viral population to treatment, confirming a loss of fitness that may have contributed to containment of the RSV infection until lymphocyte recovery. Our results have further-reaching implications for our understanding of mutagenic drugs as an approach to treating RNA-viral infection.

Data describing mutational load and mutational spectra showed that, while treatment did not clear viral infection, ribavirin reduced the fitness of the viral population via an increase in the mutation rate. Beneficial effects of mutagenic drugs in the absence of viral clearance have been observed in the treatment of persistent norovirus infection with favipiravir (Ruis et al., 2018). The clearance of infection is therefore not a necessary condition for treatment to be of clinical value, albeit that further work would be required to establish the general clinical benefit of antiviral therapies short of viral clearance. Where ribavirin was responsible for a change in viral mutation rate, favipiravir did not confer a significant additional benefit, with insufficient dosing being one possible explanation (Franco et al., 2022).

Our approach to quantifying evolutionary changes in the viral population contains multiple simplifications, working under the assumptions of a large population size and a simple population structure. In viral populations where extensive polymorphism separated viral subpopulations, more nuanced approaches to quantifying mutational load would likely be required (Raghwani et al., 2016). Measurements of viral fitness were also based on an approximation, splitting the fitness contributions from many hundreds of (presumed deleterious) mutations all across the genome from an adaptive contribution, arising from changes in composition that were characterised in terms of a few high-frequency variants. Our approach builds on previous models of virus evolution, whereby beneficial mutations compete with one another, and arise on a background of neutral and deleterious mutations (Illingworth and Mustonen, 2012; Koelle and Rasmussen, 2015; Schiffels et al., 2011). A key limitation in our modelling approach is that we were not able to characterise with precision the fitness effects of deleterious mutations. These effects are minimised at shorter viral generation times; our lower bound of 6 hours was based on previous work characterising the time from infection to the first release of viral particles from a cell during *in vitro* influenza infection (Baccam et al., 2006). Further work on within-host RSV dynamics could lead to a greater potential for evolutionary inference.

A modelling limitation to which our results are robust is the exclusion in our model of the possibility of viral compartmentalisation (Kemp et al., 2021; Sobel Leonard et al., 2017). In inferring haplotype-level fitnesses we assume a well-mixed population, with changes in viral composition occurring through competition between variants. In a case of compartmentalisation, the potential would exist for the changes we observed to have arisen in distinct parts of the airway, with a non-adaptive process of physical separation and genetic drift explaining the changes in genetic composition. In this sense, our inference of adaptive evolution is a maximal one, describing the extent of adaptation if adaptation is the sole driver of population-level change.

Different aspects of our findings are more or less generalisable to other cases of RSV infection, to other respiratory viruses and other mutagenic drugs. Under the assumption of a large population a change in viral mutation rate brought about by a specific dose of drug will lead to a consistent drop in viral fitness. Intrinsic viral fecundity may differ between patients due to differences in individualised immune responses, but the extent to which a drug interferes with replication is likely to remain constant between hosts. By contrast, changes occurring through adaptive evolution are not easily predictable, being dependent upon the precise fitness landscape (de Visser and Krug, 2014) of the genotypes close to that of the virus which founded infection. Our study of a single case shows limited viral adaptation relative to the costs imposed by mutagenesis. However, even multi-case studies face limitations. The within-host fitness landscapes of viruses can be complex and highly epistatic (Illingworth, 2015), such that even detailed characterisation of part of the genotypic space of a virus may not be informative for drawing more general conclusions.

Progress in understanding the relationship between mutagenic therapies and virus adaptation therefore remains a challenge. Mutagenesis is likely not to have a great effect where a beneficial genotype is accessible by a single mutation; the generically high mutation rate and large population sizes of RNA viruses will mean that such genotypes are created deterministically. However, mutagenesis may favour the creation of evolutionarily more distant genotypes that would otherwise not be accessible to the virus. Further, the generation of variants favoured for within-host adaptation is less of a problem if the same variants are not favoured for viral transmission (Harari et al., 2022; Lumby et al., 2018). Systematic study of within-host evolution considering multiple hosts may in future identify undesirable patterns of evolution that are promoted by antiviral mutagenesis. However, where a drug has been proven to improve short-term clinical outcomes, the evidential barrier for ceasing the use of that drug should be high.

## Methods

### Collection of sequence data

The Drugs and Therapeutics Committee at Great Ormond Street Hospital for Children approved antiviral treatment with ribavirin, nitazoxanide and favipiravir in this patient. Repeat NPAs were obtained for clinical monitoring of the viral load through polymerase chain reaction (PCR). After securing informed consent, residual viral DNA samples were collected for deep sequencing, being sequenced using the SureSelect^XT^ targeted enrichment method. Sequencing libraries were prepared using the SureSelect^XT^ low input protocol and sequenced on an Illumina MiSeq. Accompanying clinical data was retrieved from the electronic patient record and anonymised.

### Processing of sequence data

Short read data were aligned using bwa (Li and Durbin, 2010). The SAMFIRE software package was used to filter short read data, and call single- and multi-nucleotide variants from the data (Illingworth, 2016).

### Mutational load

The total mutational load was calculated as a sum of filtered variant frequencies. We note that the variant frequencies we consider in our model are very small, with the vast majority being observed at frequencies less than 1%. To this extent we carried out a filtering process, described in a previous publication (Lumby et al., 2020a), that estimates the probability of a variant of given frequency arising as the result of sequencing error; these probabilities were derived using repeat sequencing of HCV populations using the same sequencing protocol used for the data described in this study (Depledge et al., 2011).

The filtering process was conducted using a mean approach. Considering the frequency of the non-consensus nucleotide c at locus l in the genome, denoted q_lc_, we assigned the frequency-dependent probability p(q) that the variant is a false positive, defining the filtered allele frequency

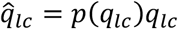

The mutational load was then calculated as

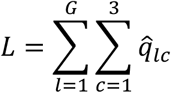

Where the sums were calculated over the length of the genome and the minority allele frequencies at each locus.

### Evolutionary model of mutational load

In a large and initially homogeneous population, mutational load increases to equilibrium according to the equation

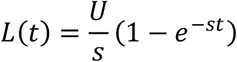

where U is the mutation rate per genome per generation, s is a constant describing the magnitude of selection against a variant, and t is time in generations (Haldane, 1937). We modified this equation to describe changes in the viral mutation rate, and variation in the magnitude of selection acting against a deleterious variant.

We modelled the change in mutational load assuming that the population begins in a situation of mutation selection balance with mutation rate U_0_ and that at specified times T_i_ the mutation rate changes so that between time T_i_ and T_i+1_ the mutation rate per genome per generation is equal to U_i_. We further assumed that the virus has a generation time of g days.

Under these circumstances, we have an estimator for the total mutational load given a constant magnitude of selection against each variant

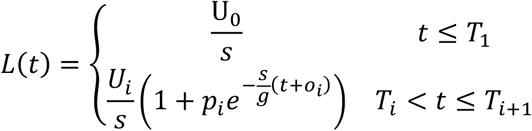

In this equation the value o_i_ is an offset term, ensuring continuity wherever the mutation rate changes. This was calculated in an iterative manner.

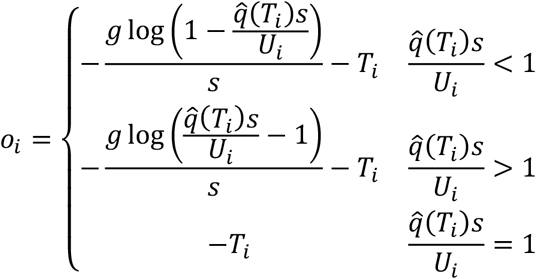

The value p_i_ controls whether the mutational load is increasing or decreasing to its new equilibrium value and was similarly calculated in an iterative manner.

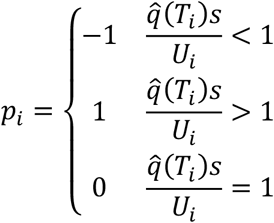

Our model contains some redundancy in so far as fixing the generation time g does not reduce the space of possible outcomes. We made the assumption that the generation time was between 6 hours, modelling the first release of influenza viruses from an infected cell (Baccam et al., 2006) and 48 hours, modelling HIV (Perelson et al., 1996). Optimal values of s and U_i_ were inferred for values of g within this range.

A preliminary optimisation of these parameters was conducted using a least squares model to fit the modelled and observed values for mutational load, before optimising the variance parameter σ to calculate the maximum likelihood.

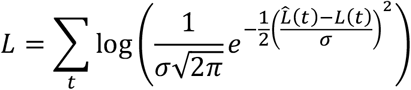

Having calculated likelihoods for models with and without changes in the mutation rate corresponding to the use of favipiravir, we compared models using the Bayesian Information Criterion (BIC).

### Calculation of changes in viral fitness due to mutational load

We used a variant of a Wright-Fisher simulation to model changes in the fitness of the viral population over time. We divide the viral population into classes according to the number of mutations in each virus. Then the value x_m_(t) describes the fraction of viruses in class m, and therefore having m mutations, after t generations.

We initialised a population in which x_0_(0) was equal to 1. In each generation mutation was modelled as a Poisson process, so that

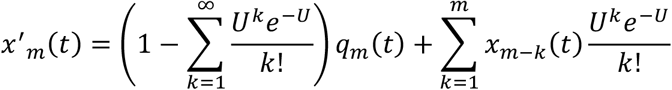

Following this selection was modelled, whereby

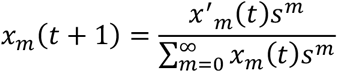

In this system the mutational load in the population was calculated as

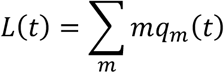

Given a model of changes in the viral mutation rate fitted to the data, we ran this simulation to numerical convergence with the mutation rate U=u_0_, and with the parameter s inferred from the model. Denoting the time t=T as the point at which treatment commenced, we altered U so that U=u_1_ for all t≥T. The fitness of the population at time t was calculated as

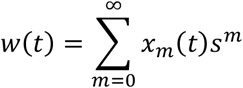

For the purpose of drawing figures linear interpolation was used to convert the statistics L(t) and w(t) from values expressed in generations to values expressed each day.

### Total evolutionary distance

The total evolutionary distance between samples was calculated as described in an earlier publication (Lumby et al., 2020a). At position l in the genome we define the vector q_l_(t) as the vector of the frequencies of the nucleotides A, C, G, and U. Given samples collected at times t_1_ and t_2_ we define the locus-specific distance d as a generalisation of the Hamming distance:

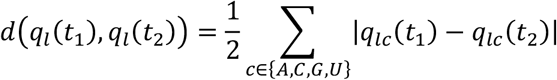

Summing these values across the genome we then calculate the total evolutionary distance

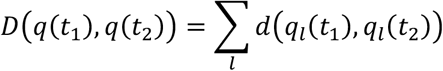

This statistic was used to evaluate the rate of evolution of the viral population, comparing how changes in D varied given greater or lesser amounts of differences between the times at which samples were collected.

### Birth-death model

A simple birth-death model was used to evaluate the potential effects of mutagenic drugs upon a viral population. Our model was described with the equation

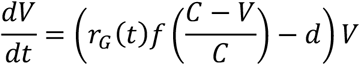

Where V is the size of the viral population, f is the pre-treatment viral fecundity, r_G_(t) is the reduction in fitness caused by the drug at time t conditional upon a generation time of G hours, d is the death rate of the virus, and C is the carrying capacity of the system. The values C=10^7^ and d=1 were chosen in this case.

### Identification of potentially selected variants

The SAMFIRE software package was used to identify potentially selected single variants in the population. Variant frequencies q(t) were identified for single nucleotide variants that reached a minor allele frequency of at least 5% in at least two samples from the population. As implemented here, this method three deterministic models of allele frequency change to the data.

1. Neutral evolution:

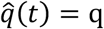

for some constant parameter q.
2. Constant selection:

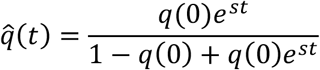

for some constant parameters q(0) and s.
3. Time-dependent selection:

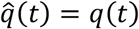

This latter model is equivalent to one in which selection is allowed to change in an unconstrained manner in between measurements of allele frequency; the result is to fit the data perfectly.

Having optimised parameters, each model was assessed using BIC. Variants for which a model of constant or time-dependent frequency is favoured over the neutral model are identified as potentially selected variants.

### Unconstrained haplotype reconstruction

Unconstrained haplotype reconstruction was carried out using a modified version of the code designed for this purpose in the VeTrans software package (Ghafari et al., 2020), extended to consider more than two samples. Given multi-locus sequence data spanning the potentially selected loci, this identified the optimum haplotype reconstruction (measured according to a likelihood fit to the data and using BIC to constrain the number of haplotypes). VeTrans is available online at www.github.com/cjri/VeTrans.

### Haplotype selection model

A constrained haplotype reconstruction was carried out using the Hapsel software package (https://github.com/cjri/Hapsel). Given multi-locus data describing changes in a population over time, plus a set of haplotype sequences H, this identifies optimal initial frequencies q_h_(0) and selection coefficients s_h_ for these haplotypes so that the evolution of the haplotype frequencies under a deterministic model best fits the observed data.

### Materials availability

Viral sequence data are available from the Sequence Read Archive with accession number PRJNA846693. Code used for the inference of parameters and for evolutionary simulation is available at https://github.com/cjri/RSVMutationalLoad. The VeTrans software package is available at https://github.com/cjri/VeTrans. The Hapsel software package is available at https://github.com/cjri/Hapsel.

### Ethics

Approval for use of the residual diagnostic specimens was obtained through the UCL/UCLH Pathogen Biobank National Research Ethics Service Committee London Fulham (Research Ethics Committee reference: 12/LO/1089).

## Data Availability

Viral sequence data are available from the Sequence Read Archive with accession number PRJNA846693.

## Acknowledgements

CJRI acknowledges UKRI Medical Research Council funding (MC_UU_12014). AYK is supported by the Wellcome Trust (222096/Z/20/Z). This work was supported in part by a Sir Henry Dale Fellowship, jointly funded by the Wellcome Trust and the Royal Society (grant numbers 101239/Z/13/Z and 101239/Z/13/A). We thank the patient, their family and the medical teams involved in their care.

## Figures

**Figure 2S1:**
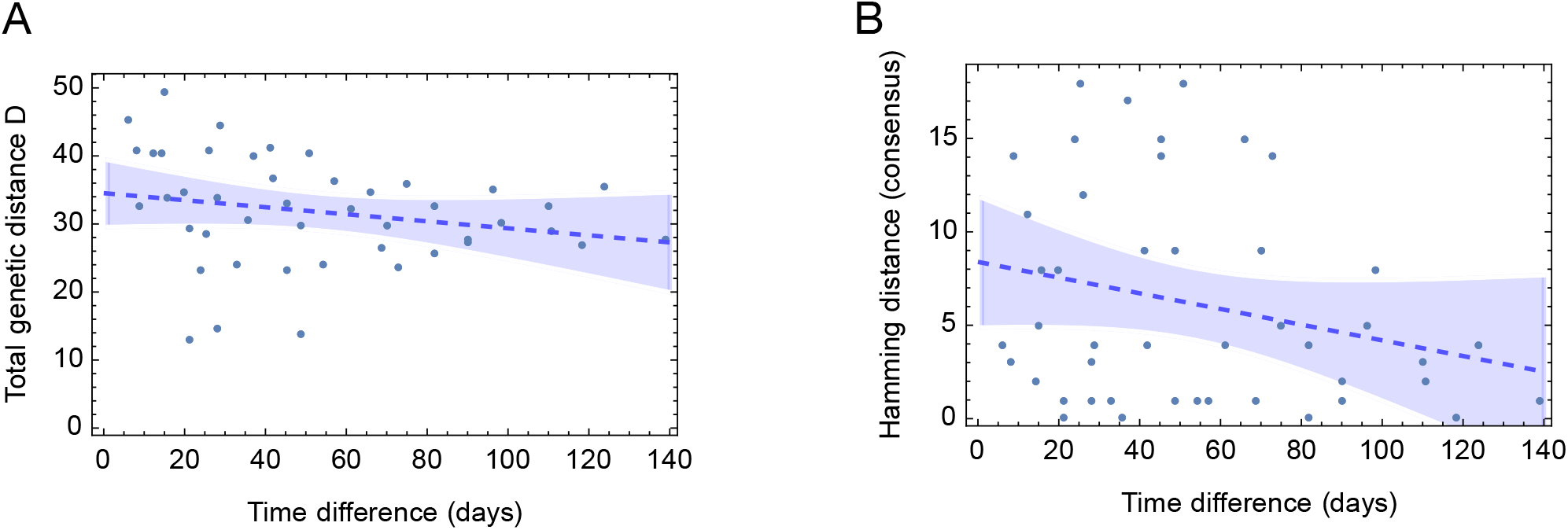
Changes in genetic distance measurements with time show little signal of consistent evolutionary change. **A**. Measurements of the total genetic distance between samples plotted against the difference in time between samples. The dashed blue line shows a best fit regression line to the data, with the shaded region showing a 95% confidence interval. The data show negligible evidence of more temporally separated samples being more genetically distinct from one another, suggesting an evolutionarily stable population. Such stability could arise either from a very large effective population size, or from evolutionary constraint, for example if all possible changes to the genetic composition of the population led to a substantial loss in population fitness. The value of the regression model at zero shows the nominal level of ‘noise’ in the sequencing process, representing the combined effects of unrepresentative sampling of a structured viral population plus errors induced in the processing and sequencing of samples. **B**. A repeat analysis, measuring the Hamming distance between consensus sequences, shows a similar result. The measure of genetic distance used in this case is less susceptible to ‘noise’.

**Figure 4S1:**
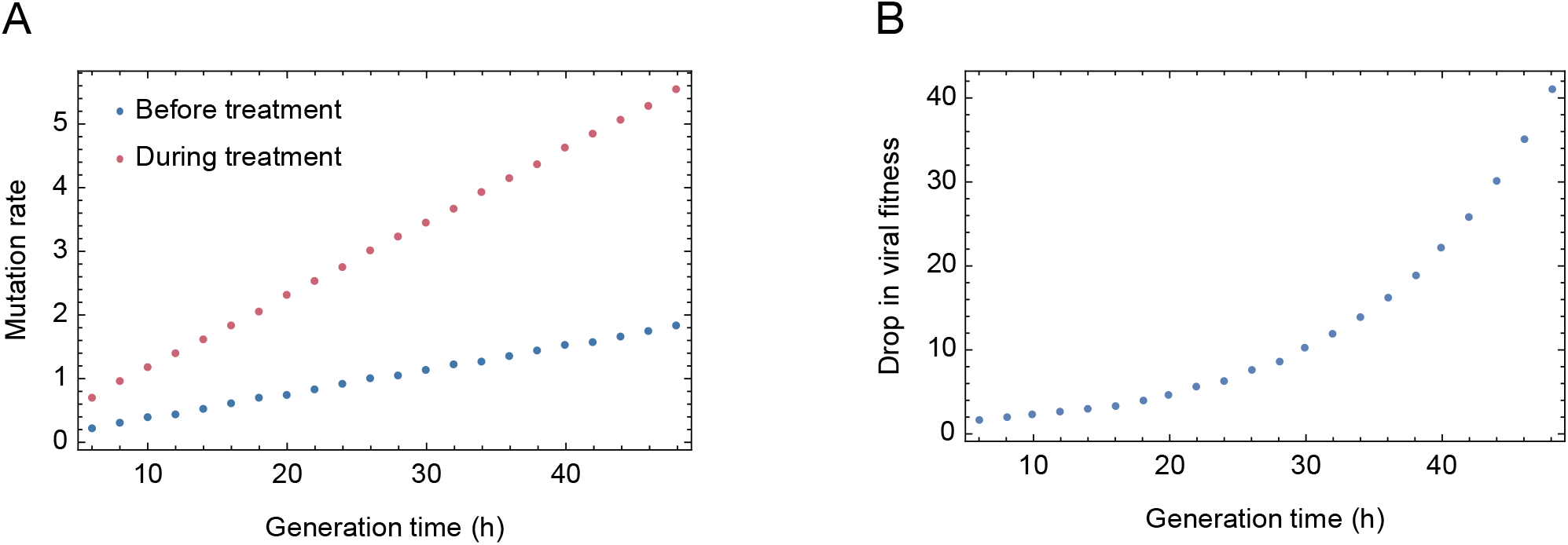
Changes in viral mutation rate and fitness under treatment. **A**. Intrinsic rates of RSV mutation inferred prior to and during treatment with ribavirin, expressed in units of mutations per genome per generation, as a function of the viral generation time in hours. **B**. Fold-drop in viral fitness as a function of the viral generation time. A drop of 10 is equivalent to a reduction to one tenth of the previous fitness.

**Figure 5S1:**
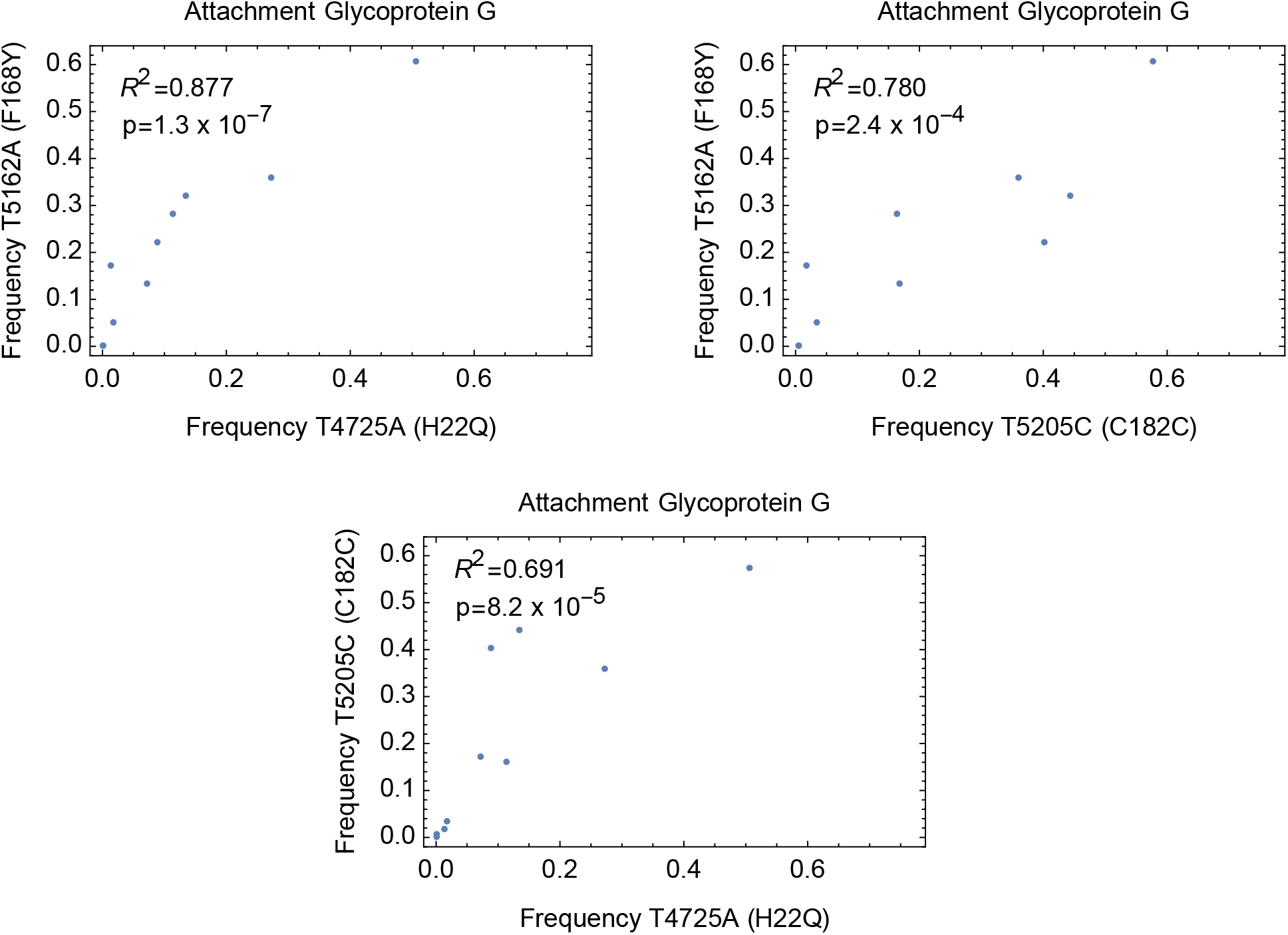
Correlations between potentially selected variants. Correlations between allele frequencies for SNPs identified as being potentially influenced by selection. Statistics show the R^2^ value from a linear regression calculation, and the p-value for a correlation test conducted using a Spearman Rank Correlation test. The pattern of frequencies is consistent with the substitution H22Q and a synonymous SNP hitchhiking with F168Y, the latter being observed at higher frequencies. Correlations shown between variants were significant at the p=0.05 level after Bonferroni correction. All such correlations occurred between SNPs within the RSV attachment glycoprotein.

**Figure 5S2:**
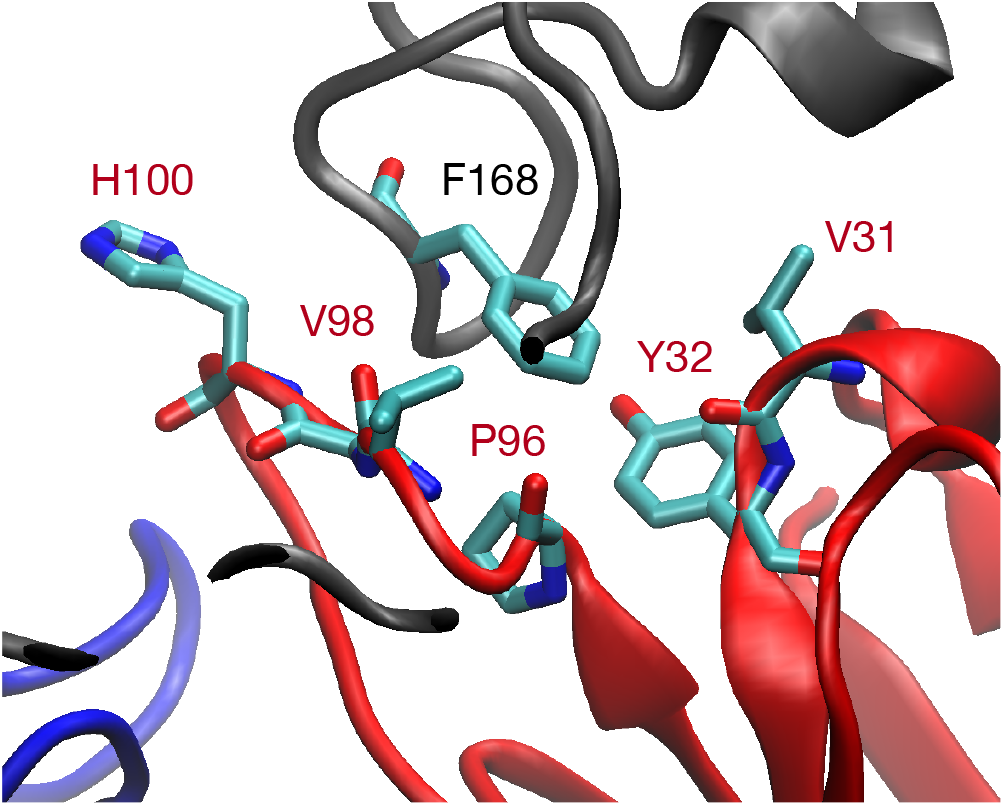
Structure of the conserved region of the G protein. The G protein is shown in gray ribbons, with the heavy and light chains of a neutralising antibody shown in blue and red. The F168Y substitution would introduce a hydroxyl group into the interface between G and the antibody, potentially disrupting antibody binding.

**Figure 5S3:**
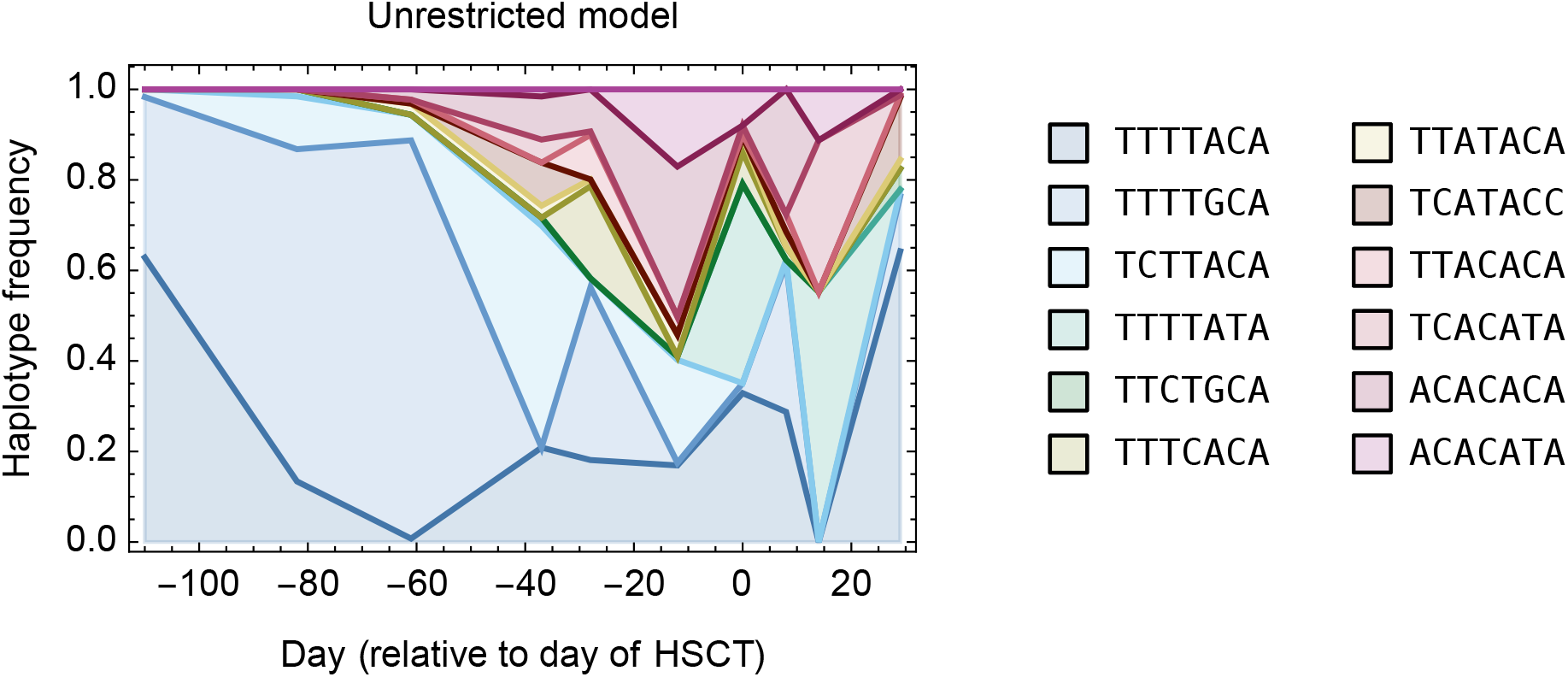
Unconstrained haplotype reconstruction describing the inferred relationships between potentially selected variants. Haplotype reconstruction was performed using multi-locus data from the seven loci at which potentially-selected variants were identified. A total of 12 haplotypes were sufficient to explain the observed sequence data across the potentially selected variant positions.

**Supporting Table 1:** Details of variant frequencies for all variants which reached a minor allele frequency of at least 5% in at least two samples. We report numbers of each nucleotide observed at each locus containing such a variant, for all time samples.

## Notes

### Competing Interest Statement

The authors have declared no competing interest.

### Author Declarations

The UCL/UCLH Pathogen Biobank National Research Ethics Service Committee London Fulham (Research Ethics Committee reference: 12/LO/1089) gave ethical approval for this work.

## References

Abdelnabi R, Foo CS, Kaptein SJF, Zhang X, Do TND, Langendries L, Vangeel L, Breuer J, Pang J, Williams R, Vergote V, Heylen E, Leyssen P, Dallmeier K, Coelmont L, Chatterjee AK, Mols R, Augustijns P, De Jonghe S, Jochmans D, Weynand B, Neyts J. 2021. The combined treatment of Molnupiravir and Favipiravir results in a potentiation of antiviral efficacy in a SARS-CoV-2 hamster infection model. eBioMedicine. doi:10.1016/j.ebiom.2021.103595

Adams R, Christenson J, Petersen F, Beatty P. 1999. Pre-emptive use of aerosolized ribavirin in the treatment of asymptomatic pediatric marrow transplant patients testing positive for RSV. Bone Marrow Transplant 24:661–664.

Baccam P, Beauchemin C, Macken CA, Hayden FG, Perelson AS. 2006. Kinetics of influenza A virus infection in humans. J Virol 80:7590–7599.

Blum VF, Cimerman S, Hunter JR, Tierno P, Lacerda A, Soeiro A, Cardoso F, Bellei NC, Maricato J, Mantovani N, Vassao M, Dias D, Galinskas J, Janini LMR, Santos-Oliveira JR, Da-Cruz AM, Diaz RS. 2021. Nitazoxanide superiority to placebo to treat moderate COVID-19 - A Pilot prove of concept randomized double-blind clinical trial. EClinicalMedicine 37:100981.

Britton PN, Hu N, Saravanos G, Shrapnel J, Davis J, Snelling T, Dalby-Payne J, Kesson AM, Wood N, Macartney K, McCullagh C, Lingam R. 2020. COVID-19 public health measures and respiratory syncytial virus. The Lancet Child & Adolescent Health. doi:10.1016/s2352-4642(20)30307-2

Brown L-AK, Freemantle N, Breuer J, Dehbi H-M, Chowdhury K, Jones G, Ikeji F, Ndoutoumou A, Santhirakumar K, Longley N, Checkley AM, Standing JF, Lowe DM. 2021. Early antiviral treatment in outpatients with COVID-19 (FLARE): a structured summary of a study protocol for a randomised controlled trial. Trials 22:193.

Bull JJ, Sanjuán R, Wilke CO. 2007. Theory of lethal mutagenesis for viruses. J Virol 81:2930– 2939.

Depledge DP, Palser AL, Watson SJ, Lai IY-C, Gray ER, Grant P, Kanda RK, Leproust E, Kellam P, Breuer J. 2011. Specific capture and whole-genome sequencing of viruses from clinical samples. PLoS One 6:e27805.

de Visser JAGM, Krug J. 2014. Empirical fitness landscapes and the predictability of evolution. Nat Rev Genet 15:480–490.

Domachowske JB, Anderson EJ, Goldstein M. 2021. The Future of Respiratory Syncytial Virus Disease Prevention and Treatment. Infect Dis Ther 10:47–60.

Franco EJ, Warfield KL, Brown AN. 2022. UV-4B potently inhibits replication of multiple SARS-CoV-2 strains in clinically relevant human cell lines. Front Biosci 27:3.

Furuta Y, Komeno T, Nakamura T. 2017. Favipiravir (T-705), a broad spectrum inhibitor of viral RNA polymerase. Proc Jpn Acad Ser B Phys Biol Sci 93:449–463.

Furuta Y, Takahashi K, Fukuda Y, Kuno M, Kamiyama T, Kozaki K, Nomura N, Egawa H, Minami S, Watanabe Y, Narita H, Shiraki K. 2002. In Vitro and In Vivo Activities of Anti-Influenza Virus Compound T-705. Antimicrobial Agents and Chemotherapy. doi:10.1128/aac.46.4.977-981.2002

Ghafari M, Lumby CK, Weissman DB, Illingworth CJR. 2020. Inferring Transmission Bottleneck Size from Viral Sequence Data Using a Novel Haplotype Reconstruction Method. J Virol 94. doi:10.1128/JVI.00014-20

Ginsburg AS, Srikantiah P. 2021. Respiratory syncytial virus: promising progress against a leading cause of pneumonia. Lancet Glob Health 9:e1644–e1645.

Goldhill DH, Langat P, Xie H, Galiano M, Miah S, Kellam P, Zambon M, Lackenby A, Barclay WS. 2019. Determining the Mutation Bias of Favipiravir in Influenza Virus Using Next-Generation Sequencing. J Virol 93. doi:10.1128/JVI.01217-18

Goldhill DH, Te Velthuis AJW, Fletcher RA, Langat P, Zambon M, Lackenby A, Barclay WS. 2018. The mechanism of resistance to favipiravir in influenza. Proc Natl Acad Sci U S A 115:11613–11618.

Gordon CJ, Tchesnokov EP, Schinazi RF, Götte M. 2021. Molnupiravir promotes SARS-CoV-2 mutagenesis via the RNA template. J Biol Chem 297:100770.

Guedj J, Piorkowski G, Jacquot F, Madelain V, Nguyen THT, Rodallec A, Gunther S, Carbonnelle C, Mentré F, Raoul H, de Lamballerie X. 2018. Antiviral efficacy of favipiravir against Ebola virus: A translational study in cynomolgus macaques. PLoS Med 15:e1002535.

Haldane JBS. 1937. The Effect of Variation of Fitness. The American Naturalist. doi:10.1086/280722

Hall CB, McBride JT, Walsh EE, Bell DM, Gala CL, Hildreth S, Ten Eyck LG, Hall WJ. 1983. Aerosolized ribavirin treatment of infants with respiratory syncytial viral infection. A randomized double-blind study. N Engl J Med 308:1443–1447.

Harari S, Tahor M, Rutsinsky N, Meijer S, Miller D, Henig O, Halutz O, Levytskyi K, Ben-Ami R, Adler A, Paran Y, Stern A. 2022. Drivers of adaptive evolution during chronic SARS-CoV-2 infections. Nat Med 28:1501–1508.

Hatter L, Eathorne A, Hills T, Bruce P, Beasley R. 2021. Respiratory syncytial virus: paying the immunity debt with interest. Lancet Child Adolesc Health 5:e44–e45.

Hayden FG, Lenk RP, Stonis L, Oldham-Creamer C, Kang LL, Epstein C. 2022. Favipiravir Treatment of Uncomplicated Influenza in Adults: Results of Two Phase 3, Randomized, Double-Blind, Placebo-Controlled Trials. J Infect Dis. doi:10.1093/infdis/jiac135

Hirsch HH, Martino R, Ward KN, Boeckh M, Einsele H, Ljungman P. 2013. Fourth European Conference on Infections in Leukaemia (ECIL-4): guidelines for diagnosis and treatment of human respiratory syncytial virus, parainfluenza virus, metapneumovirus, rhinovirus, and coronavirus. Clin Infect Dis 56:258–266.

Illingworth CJR. 2016. SAMFIRE: multi-locus variant calling for time-resolved sequence data. Bioinformatics 32:2208–2209.

Illingworth CJR. 2015. Fitness Inference from Short-Read Data: Within-Host Evolution of a Reassortant H5N1 Influenza Virus. Mol Biol Evol 32:3012–3026.

Illingworth CJR, Mustonen V. 2012. Components of selection in the evolution of the influenza virus: linkage effects beat inherent selection. PLoS Pathog 8:e1003091.

Jasenosky LD, Cadena C, Mire CE, Borisevich V, Haridas V, Ranjbar S, Nambu A, Bavari S, Soloveva V, Sadukhan S, Cassell GH, Geisbert TW, Hur S, Goldfeld AE. 2019. The FDA-Approved Oral Drug Nitazoxanide Amplifies Host Antiviral Responses and Inhibits Ebola Virus. iScience. doi:10.1016/j.isci.2019.07.003

Jayk Bernal A, Gomes da Silva MM, Musungaie DB, Kovalchuk E, Gonzalez A, Delos Reyes V, Martín-Quirós A, Caraco Y, Williams-Diaz A, Brown ML, D. J, Pedley A, Assaid C, Strizki J, Grobler JA, Shamsuddin HH, Tipping R, Wan H, Paschke A, Butterton JR, Johnson MG, De Anda C, MOVe-OUT Study Group. 2022. Molnupiravir for Oral Treatment of Covid-19 in Nonhospitalized Patients. N Engl J Med 386:509–520.

Jones HG, Ritschel T, Pascual G, Brakenhoff JPJ, Keogh E, Furmanova-Hollenstein P, Lanckacker E, Wadia JS, Gilman MSA, Williamson RA, Roymans D, van ‘t Wout AB, Langedijk JP, McLellan JS. 2018. Structural basis for recognition of the central conserved region of RSV G by neutralizing human antibodies. PLoS Pathog 14:e1006935.

Kabinger F, Stiller C, Schmitzova J, Dienemann C, Kokic G, Hillen HS, Hoebartner C, Cramer P. 2021. SARS-CoV-2 RdRp with Molnupiravir/ NHC in the template strand base-paired with A. doi:10.2210/pdb7ozu/pdb

Kaptein SJF, Jacobs S, Langendries L, Seldeslachts L, Ter Horst S, Liesenborghs L, Hens B, Vergote V, Heylen E, Barthelemy K, Maas E, De Keyzer C, Bervoets L, Rymenants J, Van Buyten T, Zhang X, Abdelnabi R, Pang J, Williams R, Thibaut HJ, Dallmeier K, Boudewijns R, Wouters J, Augustijns P, Verougstraete N, Cawthorne C, Breuer J, Solas C, Weynand B, Annaert P, Spriet I, Vande Velde G, Neyts J, Rocha-Pereira J, Delang L. 2020. Favipiravir at high doses has potent antiviral activity in SARS-CoV-2-infected hamsters, whereas hydroxychloroquine lacks activity. Proc Natl Acad Sci U S A 117:26955–26965.

Kemp SA, Collier DA, Datir RP, Ferreira IATM, Gayed S, Jahun A, Hosmillo M, Rees-Spear C, Mlcochova P, Lumb IU, Roberts DJ, Chandra A, Temperton N, CITIID-NIHR BioResource COVID-19 Collaboration, COVID-19 Genomics UK (COG-UK) Consortium, Sharrocks K, Blane E, Modis Y, Leigh KE, Briggs JAG, van Gils MJ, Smith KGC, Bradley JR, Smith C, Doffinger R, Ceron-Gutierrez L, Barcenas-Morales G, Pollock DD, Goldstein RA, Smielewska A, Skittrall JP, Gouliouris T, Goodfellow IG, Gkrania-Klotsas E, Illingworth CJR, McCoy LE, Gupta RK. 2021. SARS-CoV-2 evolution during treatment of chronic infection. Nature 592:277–282.

Koelle K, Rasmussen DA. 2015. The effects of a deleterious mutation load on patterns of influenza A/H3N2’s antigenic evolution in humans. Elife 4:e07361.

Li H, Durbin R. 2010. Fast and accurate long-read alignment with Burrows-Wheeler transform. Bioinformatics 26:589–595.

Lumby CK, Nene NR, Illingworth CJR. 2018. A novel framework for inferring parameters of transmission from viral sequence data. PLoS Genet 14:e1007718.

Lumby CK, Zhao L, Breuer J, Illingworth C Jr. 2020a. A large effective population size for established within-host influenza virus infection. Elife 9. doi:10.7554/eLife.56915

Lumby CK, Zhao L, Oporto M, Best T, Tutill H, Shah D, Veys P, Williams R, Worth A, Illingworth CJR, Breuer J. 2020b. Favipiravir and Zanamivir Cleared Infection with Influenza B in a Severely Immunocompromised Child. Clin Infect Dis 71:e191–e194.

Madhi SA, Polack FP, Piedra PA, Munoz FM, Trenholme AA, Simões EAF, Swamy GK, Agrawal S, Ahmed K, August A, Baqui AH, Calvert A, Chen J, Cho I, Cotton MF, Cutland CL, Englund JA, Fix A, Gonik B, Hammitt L, Heath PT, de Jesus JN, Jones CE, Khalil A, Kimberlin DW, Libster R, Llapur CJ, Lucero M, Pérez Marc G, Marshall HS, Masenya MS, Martinón-Torres F, Meece JK, Nolan TM, Osman A, Perrett KP, Plested JS, Richmond PC, Snape MD, Shakib JH, Shinde V, Stoney T, Thomas DN, Tita AT, Varner MW, Vatish M, Vrbicky K, Wen J, Zaman K, Zar HJ, Glenn GM, Fries LF, Prepare Study Group. 2020. Respiratory Syncytial Virus Vaccination during Pregnancy and Effects in Infants. N Engl J Med 383:426–439.

McLellan JS, Ray WC, Peeples ME. 2013. Structure and function of respiratory syncytial virus surface glycoproteins. Curr Top Microbiol Immunol 372:83–104.

Mejias A, Ramilo. 2008. Review of palivizumab in the prophylaxis of respiratory syncytial virus (RSV) in high-risk infants. Biologics: Targets & Therapy. doi:10.2147/btt.s3104

Morris SK, Dzolganovski B, Beyene J, Sung L. 2009. A meta-analysis of the effect of antibody therapy for the prevention of severe respiratory syncytial virus infection. BMC Infect Dis 9:106.

O’Brien KL, Chandran A, Weatherholtz R, Jafri HS, Griffin MP, Bellamy T, Millar EV, Jensen KM, Harris BS, Reid R, Moulton LH, Losonsky GA, Karron RA, Santosham M, Respiratory Syncytial Virus (RSV) Prevention study group. 2015. Efficacy of motavizumab for the prevention of respiratory syncytial virus disease in healthy Native American infants: a phase 3 randomised double-blind placebo-controlled trial. Lancet Infect Dis 15:1398–1408.

Ottaviano G, Lucchini G, Breuer J, Furtado-Silva JM, Lazareva A, Ciocarlie O, Elfeky R, Rao K, Amrolia PJ, Veys P, Chiesa R. 2020. Delaying haematopoietic stem cell transplantation in children with viral respiratory infections reduces transplant-related mortality. Br J Haematol 188:560–569.

Perelson AS, Neumann AU, Markowitz M, Leonard JM, Ho DD. 1996. HIV-1 dynamics in vivo: virion clearance rate, infected cell life-span, and viral generation time. Science 271:1582– 1586.

Raghwani J, Rose R, Sheridan I, Lemey P, Suchard MA, Santantonio T, Farci P, Klenerman P, Pybus OG. 2016. Exceptional Heterogeneity in Viral Evolutionary Dynamics Characterises Chronic Hepatitis C Virus Infection. PLoS Pathog 12:e1005894.

Ruis C, Brown L-AK, Roy S, Atkinson C, Williams R, Burns SO, Yara-Romero E, Jacobs M, Goldstein R, Breuer J, Lowe DM. 2018. Mutagenesis in Norovirus in Response to Favipiravir Treatment. N Engl J Med 379:2173–2176.

Schiffels S, Szöllosi GJ, Mustonen V, Lässig M. 2011. Emergent neutrality in adaptive asexual evolution. Genetics 189:1361–1375.

Schwarz G. 1978. Estimating the Dimension of a Model. The Annals of Statistics. doi:10.1214/aos/1176344136

Shi T, McAllister DA, O’Brien KL, Simoes EAF, Madhi SA, Gessner BD, Polack FP, Balsells E, Acacio S, Aguayo C, Alassani I, Ali A, Antonio M, Awasthi S, Awori JO, Azziz-Baumgartner E, Baggett HC, Baillie VL, Balmaseda A, Barahona A, Basnet S, Bassat Q, Basualdo W, Bigogo G, Bont L, Breiman RF, Brooks WA, Broor S, Bruce N, Bruden D, Buchy P, Campbell S, Carosone-Link P, Chadha M, Chipeta J, Chou M, Clara W, Cohen C, de Cuellar E, Dang D-A, Dash-Yandag B, Deloria-Knoll M, Dherani M, Eap T, Ebruke BE, Echavarria M, de Freitas Lázaro Emediato CC, Fasce RA, Feikin DR, Feng L, Gentile A, Gordon A, Goswami D, Goyet S, Groome M, Halasa N, Hirve S, Homaira N, Howie SRC, Jara J, Jroundi I, Kartasasmita CB, Khuri-Bulos N, Kotloff KL, Krishnan A, Libster R, Lopez O, Lucero MG, Lucion F, Lupisan SP, Marcone DN, McCracken JP, Mejia M, Moisi JC, Montgomery JM, Moore DP, Moraleda C, Moyes J, Munywoki P, Mutyara K, Nicol MP, Nokes DJ, Nymadawa P, da Costa Oliveira MT, Oshitani H, Pandey N, Paranhos-Baccalà G, Phillips LN, Picot VS, Rahman M, Rakoto-Andrianarivelo M, Rasmussen ZA, Rath BA, Robinson A, Romero C, Russomando G, Salimi V, Sawatwong P, Scheltema N, Schweiger B, Scott JAG, Seidenberg P, Shen K, Singleton R, Sotomayor V, Strand TA, Sutanto A, Sylla M, Tapia MD, Thamthitiwat S, Thomas ED, Tokarz R, Turner C, Venter M, Waicharoen S, Wang J, Watthanaworawit W, Yoshida L-M, Yu H, Zar HJ, Campbell H, Nair H, RSV Global Epidemiology Network. 2017. Global, regional, and national disease burden estimates of acute lower respiratory infections due to respiratory syncytial virus in young children in 2015: a systematic review and modelling study. Lancet 390:946–958.

Sobel Leonard A, McClain MT, Smith GJD, Wentworth DE, Halpin RA, Lin X, Ransier A, Stockwell TB, Das SR, Gilbert AS, Lambkin-Williams R, Ginsburg GS, Woods CW, Koelle K, Illingworth CJR. 2017. The effective rate of influenza reassortment is limited during human infection. PLoS Pathog 13:e1006203.

Sparrelid E, Ljungman P, Ekelöf-Andström E, Aschan J, Ringdén O, Winiarski J, Wåhlin B, Andersson J. 1997. Ribavirin therapy in bone marrow transplant recipients with viral respiratory tract infections. Bone Marrow Transplant 19:905–908.

Swanson KA, Settembre EC, Shaw CA, Dey AK, Rappuoli R, Mandl CW, Dormitzer PR, Carfi A. 2011. Structural basis for immunization with postfusion respiratory syncytial virus fusion F glycoprotein (RSV F) to elicit high neutralizing antibody titers. Proc Natl Acad Sci U S A 108:9619–9624.

Swanstrom R, Schinazi RF. 2022. Lethal mutagenesis as an antiviral strategy. Science 375:497–498.

Turner TL, Kopp BT, Paul G, Landgrave LC, Hayes D Jr, Thompson R. 2014. Respiratory syncytial virus: current and emerging treatment options. Clinicoecon Outcomes Res 6:217– 225.

Vanderlinden E, Vrancken B, Van Houdt J, Rajwanshi VK, Gillemot S, Andrei G, Lemey P, Naesens L. 2016. Distinct Effects of T-705 (Favipiravir) and Ribavirin on Influenza Virus Replication and Viral RNA Synthesis. Antimicrobial Agents and Chemotherapy. doi:10.1128/aac.01156-16

van Summeren J, Meijer A, Aspelund G, Casalegno JS, Erna G, Hoang U, Lina B, VRS study group in Lyon, de Lusignan S, Teirlinck AC, Thors V, Paget J. 2021. Low levels of respiratory syncytial virus activity in Europe during the 2020/21 season: what can we expect in the coming summer and autumn/winter? Euro Surveill 26. doi:10.2807/1560-7917.ES.2021.26.29.2100639

Vo NV, Young K-C, Lai MMC. 2003. Mutagenic and inhibitory effects of ribavirin on hepatitis C virus RNA polymerase. Biochemistry 42:10462–10471.

Wang Y, Fan G, Salam A, Horby P, Hayden FG, Chen C, Pan J, Zheng J, Lu B, Guo L, Wang C, Cao B. 2020a. Comparative Effectiveness of Combined Favipiravir and Oseltamivir Therapy Versus Oseltamivir Monotherapy in Critically Ill Patients With Influenza Virus Infection. The Journal of Infectious Diseases. doi:10.1093/infdis/jiz656

Wang Y, Zhong W, Salam A, Tarning J, Zhan Q, Huang J-A, Weng H, Bai C, Ren Y, Yamada K, Wang D, Guo Q, Fang Q, Tsutomu S, Zou X, Li H, Gillesen A, Castle L, Chen C, Li H, Zhen J, Lu B, Duan J, Guo L, Jiang J, Cao R, Fan G, Li J, Hayden FG, Wang C, Horby P, Cao B. 2020b. Phase 2a, open-label, dose-escalating, multi-center pharmacokinetic study of favipiravir (T-705) in combination with oseltamivir in patients with severe influenza. EBioMedicine 62:103125.

Wegzyn C, Toh LK, Notario G, Biguenet S, Unnebrink K, Park C, Makari D, Norton M. 2014. Safety and Effectiveness of Palivizumab in Children at High Risk of Serious Disease Due to Respiratory Syncytial Virus Infection: A Systematic Review. Infect Dis Ther 3:133–158.

Zhao X, Chen F-P, Sullender WM. 2004. Respiratory syncytial virus escape mutant derived in vitro resists palivizumab prophylaxis in cotton rats. Virology 318:608–612.

